# Global Landscape of Cardiovascular Research: A Continental Bibliometric Analysis Standardized by Population and Physician Density

**DOI:** 10.1101/2025.07.17.25331722

**Authors:** Mazou Temgoua, Christian Ngongang Ouankou, Walid Amara, Aimé Bonny, African CardioVAscular Research Association (ACVREA)

**Affiliations:** National College of Hospital Cardiologists, Cardio H Group, Cardiology service, Châteauroux France; Faculté de médecine de Dschang, Cameroun; Centre Hospitalier Le Raincy-Montfermeil, service de cardiologie, France; Hôpital Gynéco-obstétrique et pédiatrique de Douala, Cameroun; Faculté de médecine et des sciences pharmaceutique, Université de Douala, Cameroun

## Abstract

**Background:** Cardiovascular diseases (CVDs) remain the leading cause of global mortality. Despite rising research output, geographic disparities persist in the production and dissemination of scientific knowledge.

**Objectives:** To evaluate cardiovascular research productivity across world regions by normalizing publication output to population size and physician density, and to quantify disparities using Z-scores.

**Methods:** A comprehensive bibliometric analysis was conducted using Medline/PubMed to identify CVD-related publications across all 193 UN-recognized countries. Data were standardized using 2025 population estimates and WHO-reported physician density. Two standardized metrics were computed : publications per million population and publications per physician density. Z-scores were calculated to assess research productivity relative to global means.

**Results:** A total of 474,599 CVD publications were identified. Europe led in absolute volume (34.2%), followed by the Americas (30.7%) and Asia (25.8%). When adjusted per population density, Oceania (428.6), North America (336.7), and Western Europe (268.1) ranked highest. In contrast, Asia (15,917.3) and Sub-Saharan Africa (10,209.2) demonstrated the highest efficiency when adjusted for physician density. Z-score analysis revealed that Oceania (Z = +1.80), North America (Z = +1.25), and Western Europe (Z = +0.83) outperformed other regions in per capita productivity. However, when adjusted for physician density, Asia (Z = +2.18) and Sub-Saharan Africa (Z = +1.17) emerged as unexpectedly efficient despite limited workforce capacity. At the national level, Canada (Z = +2.44) and Japan (Z = +1.37) excelled per capita, while the United States (Z = +2.72) and Japan (Z = +1.83) led in physician-adjusted output.

**Conclusion:** Major geographic disparities exist in cardiovascular research output. Standardization by population and physician densities reveals high-efficiency zones in lowerresource settings, challenging assumptions about global scientific productivity. Strategic investment in underrepresented regions is essential to foster equitable knowledge generation and strengthen global cardiovascular health.

## Introduction

Cardiovascular diseases (CVDs) remain the leading cause of mortality worldwide, responsible for over 18 million deaths annually and representing a major burden on health systems and economies across all regions of the world (1). Despite substantial advancements in prevention, diagnosis, and management, the global cardiovascular research landscape remains highly uneven, often reflecting disparities in wealth, infrastructure, policy prioritization, and human resource allocation (2–4).

Scientific research serves as the foundation for clinical innovation, evidence-based guidelines, and public health strategies. However, a persistent imbalance exists in which individuals or institutions produce this knowledge, where it is produced, and how it is disseminated (5). Historically, high-income countries in North America and Western Europe have dominated cardiovascular research output, while low-and middle-income countries (LMICs), which bear a growing proportion of the CVD burden, remain underrepresented in scholarly contributions (6,7). Previous bibliometric studies have examined trends in global cardiovascular research output, highlighting growth in publication volume but failing to fully account for contextual variables such as population size or healthcare capacity (4,6). These dimensions are essential to understanding research equity, especially as global health initiatives increasingly emphasize capacity building and scientific inclusion (2,3,8). Standardization by population and physician density provides a more equitable lens to assess research productivity. These adjustments allow for fairer comparisons by considering both the societal burden of disease and the healthcare workforce available to engage in research (9,10). For instance, a country producing a modest number of publications may demonstrate exceptional efficiency if operating with limited human or structural resources. Conversely, high-output regions may underperform relative to their capacity when standardized for population or workforce density.

This study aims to address these gaps by conducting a comprehensive continental-level bibliometric analysis of cardiovascular publications indexed in PubMed/MedLine.This analysis was standardized by population and physician density per regions and countries in order to highlight research efficiency regardless of absolute output. This work provides insight into global imbalances in the scientific production across the continents. Our ultimate goal is to sensitize global health policies for equitable investment, and support data-driven capacitybuilding in regions historically marginalized in academic research.

## Methods

### Data Sources and Search Strategy

We conducted a structured bibliometric analysis using Medline/Pubmed, applying Boolean logic and MeSH terms (“cardiovascular diseases”, “heart diseases”, “cardiovascular”) and filtering by region through country-specific keywords. Searches were performed for total publication volume, clinical trials, and time trends (last 5 and 10 years). All 193 countries recognized by the United Nations (UN) were included in our analysis.

### Countries selection

We also randomly chose some index countries for evaluation at a national level. In each region, one of more developed and one of less developed countries were included in this analysis in the national level. Countries were selected to ensure a balanced representation across regions, income levels, healthcare capacities, and scientific output for meaningful global comparison.

### Standardization variables

We extracted :

- Total cardiovascular-related publications
- Number of clinical trials
- Publications number on the last 5 and 10 years
- Regional and countries population estimates for 2025 (Woldometer) (11)
- Physician density per 10,000 people (WHO Global Health Observatory, 2024) (12)

Two key ratios were computed: publication/population density ratio was calculated as total publication number divided by the population estimates in million people; publication/physician density ratio was the total publication number divided by the number of physicians per 10,000 inhabitants. To enable standardized comparisons of cardiovascular research productivity across regions and countries, Z-scores were calculated for two key indicators: publications per million population and publications per 10,000 physicians. For each indicator, the Z-score represents the number of standard deviations a country or region’s value deviates from the global mean. The formula used was:

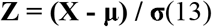

Where:

- *X* is the observed value for the country or region,
- μ is the global mean, and
- *σ* is the global standard deviation.

Z-scores were computed separately for regional and country-level analyses. For the regionallevel, global averages (across all world regions) were used as the reference for calculating Zscores. For the national-level, the reference population included all countries listed in the analysis, and Z-scores were computed based on the distribution of country-level values. This distinction was made to account for the broader heterogeneity between regions versus within countries. Higher Z-scores indicate above-average productivity relative to the global mean, while negative Z-scores indicate below-average performance.

## Results

### Regional-level Output

Europe accounted for 162,343 publications, followed by Americas (145,855), Asia (122,563), Africa (23,696), Oceania (20,142). When standardized per population density (in million people), Oceania (428.6) ranked highest in productivity followed by North America (336.7), and Western Europe (268.1). Africa and Asia were below the global median. According to physician ePiciency, Asia (15,917.3) and Sub-Saharan Africa (10,209.2) had the highest ratios whereas Western Europe (1,261.8) and North America (3,723.7) were significantly lower (Figure 1).

**Figure 1.**
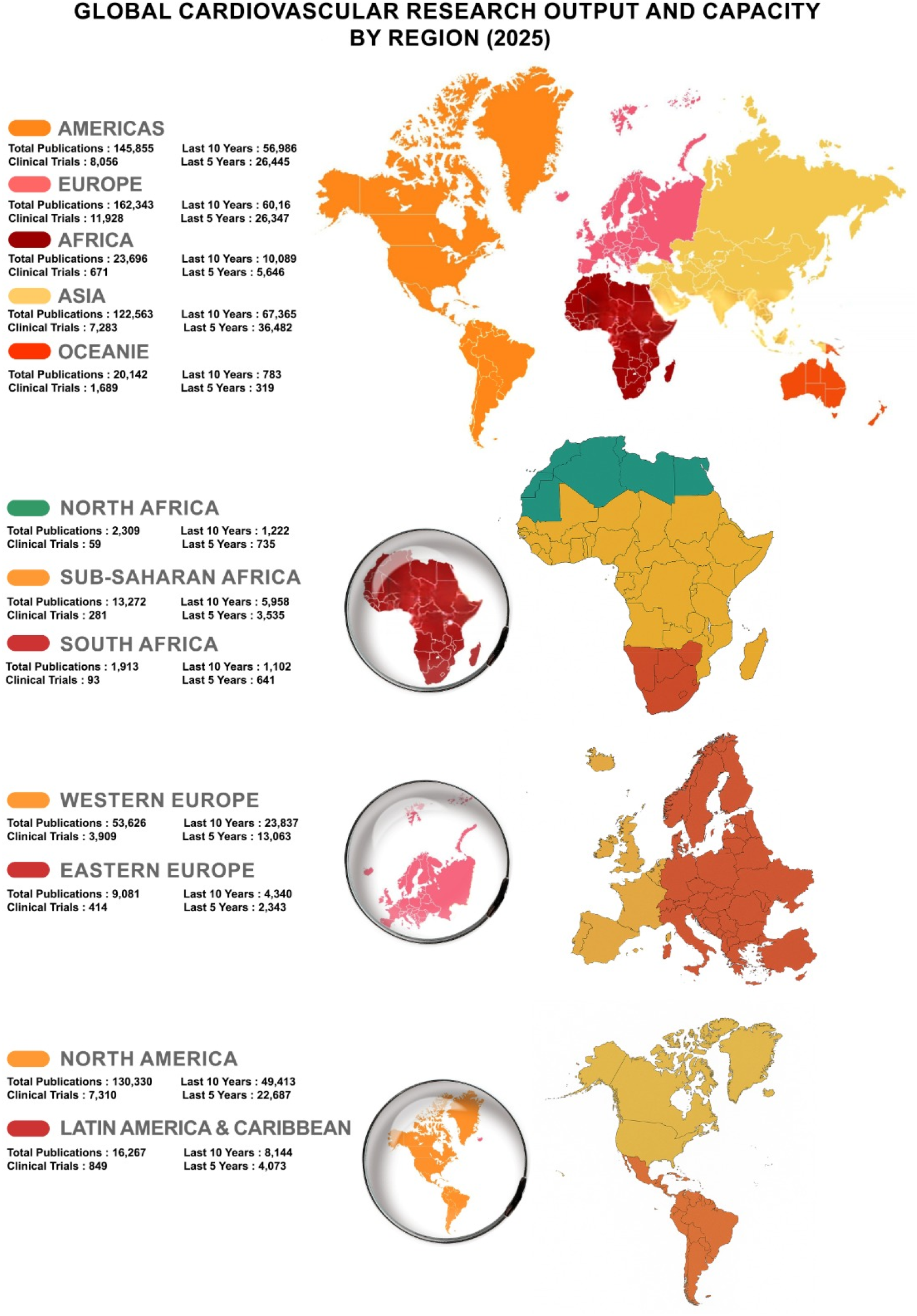
Global Distribution of Cardiovascular Research Output and Capacity by Region (2025) *Regional map of global cardiovascular research output (2025), showing total publications and clinical trials, along with trends over the last 10 and 5 years. African*

The mean of publication-to-population ratio was 129.3 (SD : 166.5), with a median of 29.4 (IQR : 243.7). For publication-to-physician density, the mean was 3795.6 (SD : 5541.2), the median was 903.7 (IQR : 3002.1). After standardization using Z-score, only North America (+1.25), Western Europe (+0.83), and Oceania (+1.80) were above average according to their population size, with Oceania leading significantly. In contrast, Asia (+2.18) and Sub-Saharan Africa (+1.17) showed exceptionally high ePiciency when adjusted by physician density (Table 1).

**Table 1.** Standardized Cardiovascular Research Productivity by Region (Z-Scores, 2025)

| Region | Population (Million people) | Physician Density<br>(Per 10 000 inhabitants) | Publications / Population Density | Publications / Physician Density | Z-score/ Population Density | Z-score/ Physician Density |
| --- | --- | --- | --- | --- | --- | --- |
| North Africa | 275 | 3.2 | 8.4 | 721.6 | -0.73 | -0.56 |
| Sub-Saharan Africa* | 1,200 | 1.3 | 11.1 | 10,209.2 | -0.71 | +1.17 |
| South Africa | 65 | 7.9 | 29.4 | 242.1 | -0.60 | -0.64 |
| Asia | 4,828 | 7.7 | 25.4 | 15,917.3 | -0.62 | +2.18 |
| Western Europe | 200 | 42.5 | 268.1 | 1,261.8 | 0.83 | -0.46 |
| Eastern Europe | 285 | 29.0 | 31.9 | 313.1 | -0.59 | -0.63 |
| North America | 387 | 35.0 | 336.7 | 3,723.7 | 1.25 | -0.01 |
| Latin America & Caribbean | 667 | 18.0 | 24.4 | 903.7 | -0.63 | -0.52 |
| Oceania | 47 | 23.2 | 428.6 | 868.2 | 1.80 | -0.53 |

*\*All Sub-Saharan African countries were included except South Africa, which was analyzed separately to reduce regional disparities.*

### National-level Output

Country-specific analysis showed several high performers : Japan (126.9 publications/million people; 589.2 per 10 000 physicians), USA (83.5/million people; 787.6 per 10 000 physicians), Canada (187.8/million people; 266.4 per 10 000 physicians). South Africa also showeded high relative output despite low physician density (7.9/10 000 physicians, Table 2).

**Table 2.** Cardiovascular Research Output and Capacity by Country (2025)

| **Countries** | **Total Publications** | **Last 10 Years** | **Last 5 Years** | **Clinical Trials** | **Population Density**<br>**(per million people)** | **Physician Density**<br>**(Per 10 000 inhabitants)** | **Publications / Population Density** | **Publications / Physician Density** |
| --- | --- | --- | --- | --- | --- | --- | --- | --- |
| USA | 28,990 | 14,565 | 8,554 | 1,392 | 347 | 36.81 | 83.5 | 787.6 |
| Canada | 7,511 | 3,730 | 2,015 | 731 | 40 | 28.19 | 187.8 | 266.4 |
| Brazil | 4,974 | 2,796 | 1,562 | 237 | 213 | 23.57 | 23.35 | 211.03 |
| Colombia | 786 | 557 | 340 | 25 | 53 | 25.35 | 14.8 | 31.0 |
| Germany | 8,506 | 3,865 | 2,078 | 752 | 84 | 45.34 | 101.26 | 187.6 |
| Greece | 1215 | 532 | 293 | 45 | 10 | 65.76 | 121.5 | 18.5 |
| Russia | 1248 | 618 | 295 | 104 | 144 | 51.11 | 8.7 | 24.4 |
| Bulgaria | 212 | 100 | 52 | 12 | 6,7 | 43.28 | 31.64 | 4.9 |
| Japan | 15,608 | 7,286 | 4,239 | 1049 | 123 | 26.49 | 126.9 | 589.2 |
| Pakistan | 1,471 | 954 | 564 | 87 | 254 | 11.6 | 5.8 | 126.8 |
| Malaysia | 1032 | 629 | 380 | 48 | 36 | 23.41 | 28.7 | 44.08 |
| Egypt | 683 | 404 | 249 | 41 | 118 | 6.71 | 5.8 | 101.8 |
| Niger | 102 | 34 | 20 | 5 | 28 | 0.38 | 3.6 | 268.4 |
| Burundi | 11 | 8 | 6 | 0 | 14 | 0.78 | 0.79 | 14.1 |
| Botswana | 84 | 64 | 35 | 3 | 2,5 | 3.79 | 22.6 | 22.2 |
| South Africa | 1913 | 1102 | 641 | 93 | 65 | 7.94 | 29.4 | 240.9 |

Across countries, the mean number of publications per million people was 49.8 (SD : 56.5), with a median of 26.0 (IQR : 79.96). Concerning publications per physician density, the mean was 183.7 (SD : 222.0), the median was 114.3 (IQR : 223.4). According to their population size, Canada (z = +2.44) and Japan (z = +1.37) are the most outstanding performers. Countries such as Burundi, Niger, Egypt, and Pakistan showed very low publication rates per capita (z < −0.75). According to the density of physicians, the USA (z = +2.72) and Japan (z = +1.83) stand out as producing the highest number of publications. Bulgaria, Greece, Russia, Botswana, and Burundi have significantly low z-scores (z < −0.7). Table 3.

**Table 3.** Standardized Cardiovascular Research Productivity by Country (Z-Scores, 2025)

| **Country** | **Z-score/**<br>**Population Density** | **Z-score/**<br>**Physician Density** |
| --- | --- | --- |
| USA | 0.60 | 2.72 |
| Canada | 2.44 | 0.37 |
| Brazil | -0.47 | 0.12 |
| Colombia | -0.62 | -0.69 |
| Germany | 0.91 | 0.02 |
| Greece | 1.27 | -0.74 |
| Russia | -0.73 | -0.72 |
| Bulgaria | -0.32 | -0.81 |
| Japan | 1.37 | 1.83 |
| Pakistan | -0.78 | -0.26 |
| Malaysia | -0.37 | -0.63 |
| Egypt | -0.78 | -0.37 |
| Niger | -0.82 | 0.38 |
| Burundi | -0.87 | -0.76 |
| Botswana | -0.48 | -0.73 |
| South Africa | -0.36 | 0.26 |

## Discussion

This global bibliometric assessment reveals striking disparities in cardiovascular research productivity when analyzed both in absolute terms and adjusted for regional and national capacities. While Western Europe, North America, and Oceania unsurprisingly dominate per capita output, the relative efficiency in Asia and Sub-Saharan Africa when standardized by physician density is a critical finding that challenges conventional narratives around research equity. Asia’s performance, particularly in countries like Japan and China reflects national strategies emphasizing biomedical innovation, infrastructure investment, and strong academic cultures (14–16). These countries often benefit from centralized research funding and prioritized Non-communicable diseases (NCD) research in their public health agendas. In contrast, Sub-Saharan Africa’s (SSA) high publication-to-physician density ratio is likely the result of both a low baseline number of physicians and the growing contribution of emerging centers of excellence. These findings should be interpreted cautiously: while relative efficiency is high, absolute output and broader capacity for implementation remain limited (17,18).

### Barriers for research outputs in Africa

Our study showeded the rate of less than 5% research output from Africa for than 18% of world population. Critical barriers in SSA include the absence of formal research curricula in most medical schools, a lack of institutional incentives for clinician-scientists, lack of a dedicated research team, and the economic precarity of physicians who are more preoccupied with making ends meet than pursuing unfunded research (19,20). The situation is compounded by a near-total absence of national funding structures. There are often no competitive calls for research proposals from government institutions, and university research budgets lack transparency and accountability. These factors collectively contribute to a research environment where talent is abundant but structurally under-supported. As a result, many highly-skilled African researchers and clinicians migrate to countries where scientific infrastructures are more favorable and career prospects more sustainable. Several recent studies demonstrate that this emigration enhances scientific productivity in developed countries through knowledge spillovers, professional networks, and increased research output while simultaneously reducing research capacity and productivity in countries of origin due to the depletion of human capital (21–23). This brain drain further deepens the global imbalance in research equity and innovation.

### Competitive research workflow in developed regions

North America and Western Europe, despite producing the majority of high-impact research and clinical guidelines, showed lower physician-standardized outputs. This may reflect the higher clinical workload, broader specialization, or institutional separation between clinical and research responsibilities (3,5,24).

### Barriers for research outputs in Latin America and Eastern Europe

Latin America and Eastern Europe appear underrepresented in both absolute and standardized terms. Barriers may include limited research funding, fewer collaborative networks, language barriers, and lower visibility in international databases (25).

### Disparities at national level

There is marked heterogeneity across countries illustrating global imbalances. Canada and Japan, for instance, demonstrated high productivity both in per capita terms (Z = +2.44 and +1.37, respectively) and when adjusted for physician density (Z = +1.83 for Japan). This dual efficiency underscores well-integrated research ecosystems supported by long-standing academic traditions and stable funding environments (15). The United States also stands out for its high physician-standardized output (Z = +2.72), despite lower rankings in per capita terms, possibly reflecting intensive institutional support for research within large academic health systems (26). Conversely, African countries such as Burundi, Niger, and Egypt as well as Pakistan in Asia exhibited some of the lowest Z-scores in population-adjusted output (Z < –0.75), reflecting structural constraints such as limited research infrastructure, economic hardship, and health workforce shortages (9). In some Eastern European including Bulgaria, Greece, Russia, Z-scores were also markedly low when adjusted for physician density, suggesting a disconnect between available workforce and research engagement. The disparity in research funding across Europe is well known. Despite substantial investment from the European Union to support research initiatives, countries in Eastern Europe remain largely underrepresented among funding recipients. In the field of cardiovascular research in particular, there is a paradoxical trend whereby regions with the highest mortality rates receive the least financial support. For instance, during the early 2010s, no major grants exceeding €100,000 per project were awarded to institutions in several Eastern European countries, highlighting a persistent imbalance in resource allocation (27).

From a global health equity perspective, these findings have profound implications. International initiatives like the WHO NCD Action Plan (28) and UNESCO Open Science framework (29) emphasize the democratization of knowledge generation. Supporting regional research capacity through funding, partnerships, and inclusive publishing practices are essential to close the cardiovascular knowledge gap. Lastly, the metrics used here particularly normalization by physician density offer a novel perspective on scientific efficiency. However, they must be triangulated with data on research quality, citation impact, and implementation outcomes in future studies.

### Study limitations

This study has several limitations. First, PubMed indexing may omit local or regional journals, potentially underestimating contributions from low- and middle-income countries. Second, publication counts do not reflect research quality, clinical impact, or citation influence, which are important dimensions of scientific productivity. Third, country affiliations were based on title and abstract data and may not fully capture cross-border collaborations or the contribution of diaspora researchers. Additionally, the use of only three broad MeSH terms may have led to the inclusion of articles not exclusively focused on cardiovascular diseases. The completeness and accuracy of physician density data from the WHO Global Health Observatory may vary between countries, possibly affecting standardization precision. The analysis did not distinguish between physician and non-physician authors, which limits the interpretation of scientific output relative to clinical workforce capacity. Finally, while Z-score normalization offers a useful comparative metric, it may be less intuitive for non-specialist readers and should be interpreted alongside absolute values.

Despite these limitations, this study provides one of the first standardized, continent-wide assessments of cardiovascular research productivity, accounting for both population size and physician density. By highlighting hidden efficiencies and structural imbalances, it offers actionable insights for global health funders, policymakers, and academic leaders seeking to promote research equity and strengthen capacity in underrepresented regions.

## Conclusion

While global cardiovascular research output is increasing, it remains concentrated in high-income settings. Adjusting for local capacity (population and physician workforce) uncovers high-ePiciency zones in lower-resource areas. Prioritizing equity-driven research investments is essential for addressing the global burden of CVDs and reverse their worsening trajectory.

## Data Availability

All data produced in the present study are available upon reasonable request to the authors

## Acknowlegment

we are deeply greatful to Ms Vicky NLIBA for the assistance that she provided during the process of the designing and writting the manuscript.

## Abbreviations

CVDs: Cardiovascular Diseases
GBD: Global Burden of Disease
IQR: Interquartile Range
LMICs: Low-and Middle-Income Countries
MeSH: Medical Subject Headings
NCDs: Non-Communicable Diseases
SD: Standard deviation
SSA: Sub-Saharan Africa
UNESCO: United Nations Educational, Scientific and Cultural Organization
USA: United States of America
WHO: World Health Organization
Z: Z-score μ = Mean
σ: Standard Deviation

## Author Contributions

***Study conception* :** Mazou Temgoua (MT), Aimé Bonny (AB).

***Data collection* :** Mazou Temgoua (MT), Christian Ngongang Ouankou (CNO).

***Critical revision and final approval of the manuscript* :** Mazou Temgoua (MT), Aimé Bonny (AB), Christian Ngongang Ouankou (CNO), Walid Amara (WA).

